# LifeLine.ai: A SigLIP-MedGemma Multimodal Framework with KTO Alignment for Open-Vocabulary 12-Lead ECG Report Generation

**DOI:** 10.64898/2026.11.25.26368934

**Authors:** Asad Irfan, Khurram Khan Jadoon, Muhammad Hanif

## Abstract

Automated twelve-lead electrocardiogram (ECG) interpretation has historically relied on discriminative classifiers limited to closed-set diagnostic labels, signal reconstruction, or saliency heatmaps. While categorizing predefined arrhythmias well, these static classifiers cannot generate narrative clinical reports, incorporate patient history, or support interactive clinical question-answering (Q&A). We present LifeLine.ai, an end-to-end multimodal Vision-ECG-Language framework for automated cardiac diagnosis and clinical reporting. After early attempts at waveform peak-tracking and coordinate mapping failed against complex morphological variations, we leveraged large-scale PhysioNet repositories (MIMIC-IV and PTB-XL). Because hardware limits precluded training on over 800,000 raw records, we curated a balanced subset to estimate performance, converting one-dimensional signals into 3×4 twelve-lead grids with continuous Lead II rhythm strips at the bottom. Our pipeline integrates YOLOv11s localization, a LoRA-adapted Google SigLIP encoder, a trainable Multi-Layer Perceptron projector, and a 4-bit quantized MedGemma-4B model, complemented by a separate Kahneman-Tversky Optimization (KTO) pipeline built on collected user feedback and a multi-LLM consensus engine. On this compact subset, SigLIP outperformed Swin Transformer encodings (∼0.21 vs. ∼0.25 validation loss), achieving 71.0% open-vocabulary precision and a 75.0% contextual F1 score across a cloud deployment spanning Vercel, mobile apps, Amazon EC2, Firebase, Hugging Face, Modal GPUs, and “lifelinecg-sdk”. Our code is available at https://github.com/Asad939asad/LifeLine.ai-.

## 1. INTRODUCTION

Cardiovascular diseases remain the leading cause of global mortality, making the twelve-lead electrocardiogram (ECG) a primary non-invasive tool in emergency medicine and routine cardiology. In clinical practice, cardiologists do not reduce an ECG to a single classification tag. Instead, they interpret various morphological patterns across twelve spatial leads and a continuous rhythm strip, assessing P-wave progression, QRS duration, ST-segment deviations, and T-wave changes, alongside patient history to produce a narrative report that explains the underlying mechanism, differential diagnoses, and urgency of care. Clinical decision-making is also interactive, requiring follow-up question-answering (Q&A) on specific waveform anomalies and triage steps.

Most automated ECG systems, however, frame interpretation as closed-set classification. Discriminative CNNs (*ECG-Image* [1]), ensemble models (*xECGArch* [3]), and hybrid digitization pipelines (*Digitized-ECG* [2]) map clean digital plots or reconstructed 1D signals to fixed diagnostic labels. While effective on narrow label sets, these systems face four practical limitations: (1) they cannot generate narrative clinical reasoning, (2) they lack a language backbone for interactive Q&A, (3) post-hoc saliency overlays (e.g., Grad-CAM or Deep Taylor Decomposition) highlight spatial regions without explaining findings in medical terms, and (4) they often break on real-world photos of thermal ECG printouts containing background clutter, skew, or grid noise.

Our initial attempts to address this problem using rule-based peak tracking (P, Q, R, S, and T-wave coordinate extraction) proved brittle under baseline wander, lead overlaps, and pathological waveform distortions. We therefore pivoted to a data-driven Vision-Language Model (VLM) approach built on raw 1D ECG repositories from PhysioNet (PTB-XL and MIMIC-IV, spanning over 800,000 records). Because training on the full raw archive is infeasible on single-GPU academic hardware (NVIDIA RTX A2000, 12 GB VRAM) and suffers from severe majority-class skew, we curated a class-balanced subset of approximately 50000 samples and built a signal-to-image synthesis pipeline that renders raw waveforms into standardized 3×4 twelve-lead grids with continuous Lead II strips paired with structured JSON clinical narratives.

To bridge ECG morphology and clinical language within 12 GB VRAM, we developed a QLoRA fine-tuning pipeline around a 4-bit quantized MedGemma-4B backbone and compared vision encoders directly. Pairing MedGemma-4B with a windowed Swin Transformer fragmented horizontal waveform continuity across local patches, plateauing at ∼0.25 validation loss. Replacing Swin with a LoRA-adapted Google SigLIP encoder connected via a trainable Multi-Layer Perceptron (MLP) projector preserved cross-lead continuity and lowered validation loss to ∼0.21. To further align narrative quality with user expectations without costly pairwise preference labeling, we added a separate Kahneman-Tversky Optimization (KTO) alignment stage trained on binary user feedback, toggling LoRA adapters to evaluate the frozen reference policy while keeping the MLP projector fixed. Integrated with Real-ESRGAN enhancement, padding-aware YOLOv11s localization, and a multi-LLM consensus layer, LifeLine.ai delivers end-to-end open-vocabulary ECG interpretation in the cloud.

### Main Contributions

### 1.1. Class-Balanced 3×4 ECG Synthesis Pipeline

A signal-to-image pipeline converting raw PhysioNet (PTB-XL and MIMIC-IV) 1D waveforms into standardized 3×4 twelve-lead grids with continuous Lead II rhythm strips and structured JSON clinical narratives, balancing rare arrhythmias against dominant classes (NORM, IMI, ASMI, LVH).

### 1.2. Vision Encoder Ablation & KTO Preference Alignment on MedGemma-4B

An end-to-end trainable SigLIP-to-MedGemma MLP projection architecture that outperforms windowed Swin encodings (∼0.21 vs. ∼0.25 validation loss), paired with a standalone KTO stage using binary user feedback and adapter-toggling reference computation.

### 1.3. Memory-Efficient Single-GPU Training & Open-Vocabulary Evaluation

A reproducible 4-bit NF4 QLoRA recipe (8-bit AdamW, effective batch size 32, cosine schedule, target-only loss masking) trained on a single 12 GB RTX A2000 GPU, achieving 71.0% open-vocabulary precision and 75.0% contextual F1 on unconstrained clinical report generation and Q&A.

### 1.4. Full-Stack Cloud Deployment & SDK (LifeLine.ai)

A hardware-agnostic inference pipeline (Real-ESRGAN, YOLOv11s, fine-tuned VLM, and multi-LLM consensus) deployed across web/mobile clients, EC2/Firebase middleware, Hugging Face FastAPI endpoints, Modal serverless GPUs, and the open-source lifelinecg-sdk Python package.

## 2. RELATED WORK

### A. Image-Based and Digitized ECG Analysis

Early automated electrocardiogram (ECG) classification systems relied predominantly on clean one-dimensional digital voltage signals, as demonstrated at scale by Ribeiro et al. [15] using residual neural networks across more than two million 12-lead exams. However, reliance on raw signal files restricts clinical deployment where records exist only as paper printouts or scanned images. To handle image-based inputs, Sangha et al. [16] and ECG-Image [1] applied convolutional neural networks (CNNs) directly to plotted 12-lead ECG images, while ECG-Image-Kit [17] introduced synthetic artifact generation to support paper-to-signal digitization benchmarks. Along similar lines, Digitized-ECG [2] proposed a hybrid digitization workflow that reconstructs one-dimensional time-series waveforms from plots before classifying arrhythmias, and xECGArch [3] combined independent short-term and long-term CNN branches in an ensemble architecture to capture morphological and rhythm-level cardiac features for atrial fibrillation detection. Although these methods achieve high classification accuracy on predefined benchmarks, they share fundamental structural limitations. First, they operate as closed-set discriminative classifiers or signal digitizers restricted to fixed diagnostic taxonomies. Second, their interpretability relies on post-hoc visual attribution such as Grad-CAM saliency overlays in [1], [16] or Deep Taylor Decomposition in xECGArch [3], which highlight spatial pixel regions without providing clinical reasoning in natural language. Consequently, none of these approaches generate narrative clinical reports, incorporate patient-specific context, or support interactive clinical question-answering (Q&A).

### B. Medical Vision-Language Models

Multimodal Vision-Language Models (VLMs) align visual representations with generative language models, enabling open-vocabulary reporting and interactive diagnostic reasoning. Broad biomedical foundation models such as Med-PaLM M [18] and BiomedCLIP [19] demonstrated strong cross-modal transfer across clinical imaging and text tasks, while domain-specific ECG models such as PULSE [20] showed that instruction-tuning multimodal LLMs on rendered ECG images improves diagnostic interpretation over general-purpose models though typically at the cost of million-scale instruction datasets and multi-GPU cluster training. More compactly, MedGemma [4] builds upon the Gemma 3 architecture through medical-domain pre-training, reducing clinical hallucinations relative to general-purpose language models. However, selecting a suitable vision encoder for dense 12-lead ECG plots remains critical. Hierarchical architectures such as the Swin Transformer [5] restrict self-attention to non-overlapping local windows before merging patches across stages. While effective for natural images, windowed partitioning fragments continuous horizontal ECG waveforms and disrupts cross-lead temporal coherence across 3×4 grid layouts. In contrast, SigLIP [6] replaces softmax-normalized contrastive training with a pairwise sigmoid loss, producing unified patch embeddings across the entire image. Preserving continuous waveform morphology across all twelve leads and the rhythm strip enables more consistent cross-modal alignment with clinical text than fragmented local window encodings.

### C. Efficient Fine-Tuning and Preference Alignment

Adapting multi-billion-parameter VLMs to specialized biomedical tasks under single-GPU memory constraints requires parameter-efficient training. Low-Rank Adaptation (LoRA) [12] freezes pre-trained weights and injects trainable low-rank decomposition matrices into transformer projections, drastically reducing trainable parameters. Building on this, QLoRA [7] further lowers memory overhead by quantizing frozen base weights to 4-bit NormalFloat (NF4) precision with double quantization while optimizing low-rank adapters in higher precision, enabling fine-tuning on commodity GPUs. Beyond supervised fine-tuning, generated clinical descriptions must align with human expectations for diagnostic clarity and accuracy. Reinforcement learning from human feedback (RLHF) [21] aligns language models via explicit reward modeling and policy optimization, while Direct Preference Optimization (DPO) [22] simplifies alignment into a closed-form objective without a separate reward model. However, both RLHF and DPO require paired preference datasets containing both chosen and rejected outputs for each prompt, which are costly and difficult to collect in medical settings. Kahneman-Tversky Optimization (KTO) [8] eliminates this pairing requirement by framing alignment as prospect-theoretic utility optimization over standalone binary desirability labels. Consequently, KTO allows medical VLMs to be aligned directly from binary user feedback signals collected during system evaluation.

## 3. DATASET AND VISUAL PREPROCESSING PIPELINE

### A. Data Sources and Class-Balanced Curation

Raw electrocardiography records were aggregated from two PhysioNet repositories: PTB-XL [9] and MIMIC-IV-ECG [10], collectively spanning over 800,000 one-dimensional waveform recordings paired with cardiologist reports and structured metadata. For MIMIC-IV-ECG, study metadata, automated machine measurements (PR/QRS/QT intervals, axis parameters, and sequential text fields), and clinical note links were merged on study_id, deduplicating repeated notes and removing records with missing waveform paths.

Training a multimodal vision-language model across all 800,000 records is infeasible on a single NVIDIA RTX A2000 GPU (12 GB VRAM) due to memory, I/O, and training-time limits. Rather than taking a naive random subsample, which would be heavily skewed toward common conditions across PTB-XL’s 71 SCP diagnostic statements, namely normal sinus rhythm (NORM), inferior myocardial infarction (IMI), anteroseptal myocardial infarction (ASMI), and left ventricular hypertrophy (LVH). We paired dataset reduction with targeted class balancing to curate a compact corpus of approximately 50,000 diverse ECG records. Specifically, Stage 1 alignment used a 97% - 3% train/validation split across valid records, while Stage 2 retained 100% of rare-pathology recordings and downsampled studies whose active labels fell exclusively within {NORM, IMI, ASMI, LVH} to 5% (p = 0.05), split 95% - 5% for training and validation. This strategy reduced total training volume to fit single-GPU constraints while preventing majority classes from dominating gradient updates.

### B. 1D Waveform to 3×4 Visual Grid Synthesis and Inference Sanitization

Because PhysioNet distributes raw voltage arrays rather than clinical charts, we built a parallelized WFDB rendering pipeline that reads paired binary (.dat) and header (.hea) files at each record’s native sampling rate (100 Hz for PTB-XL; native header rates for MIMIC-IV-ECG) and discards NaN or flat zero-variance channels. For each study, the script slices 4-second windows across all 12 standard leads (I, II, III, aVR, aVL, aVF, V1–V6) and a continuous 10-second Lead II rhythm strip (aligned at the bottom). Short leads are rendered at 150 DPI on 5.5 x 0.9-inch canvases (825 x 135 px) and the rhythm strip on a 9.5 x 1.0-inch canvas (1425 x 150 px) over a white background overlaid with semi-transparent pink grid lines at 5.6-pixel intervals. As shown in Fig. 2, these panels compose a standard 3×4 clinical grid resized to 224 x 224 RGB pixels for vision encoding. During 13-view vision pre-training, absent channels are padded with blank frames and augmented with brightness/contrast jitter (0.2), random sharpness (factor 2, p = 0.5), and mild affine perturbations (2° rotation, 0.02 translation) to prevent overfitting to grid color.

**Fig 1.**
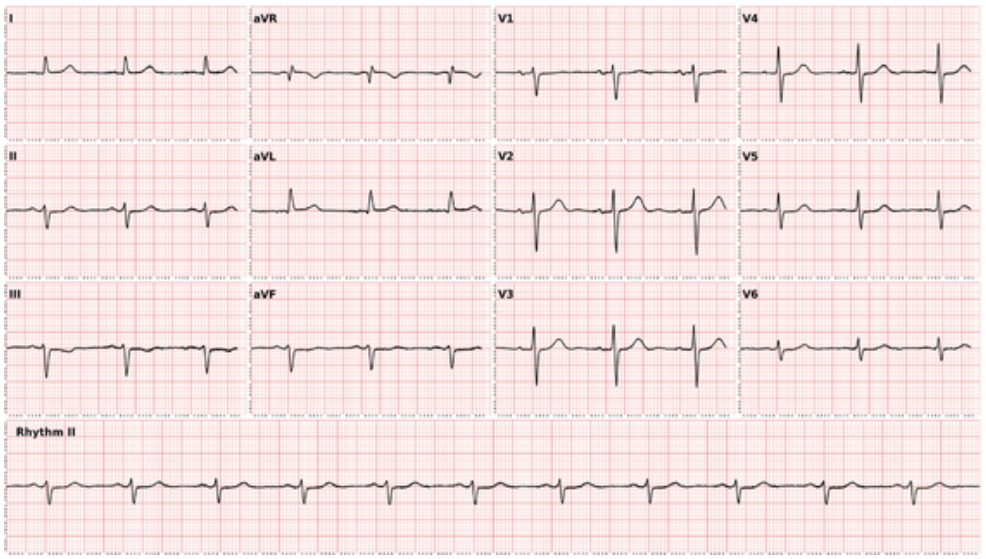
3×4 ECG image constructed from a 1D ECG recording, with an extended Lead II rhythm strip at the end.

**Fig 2.**
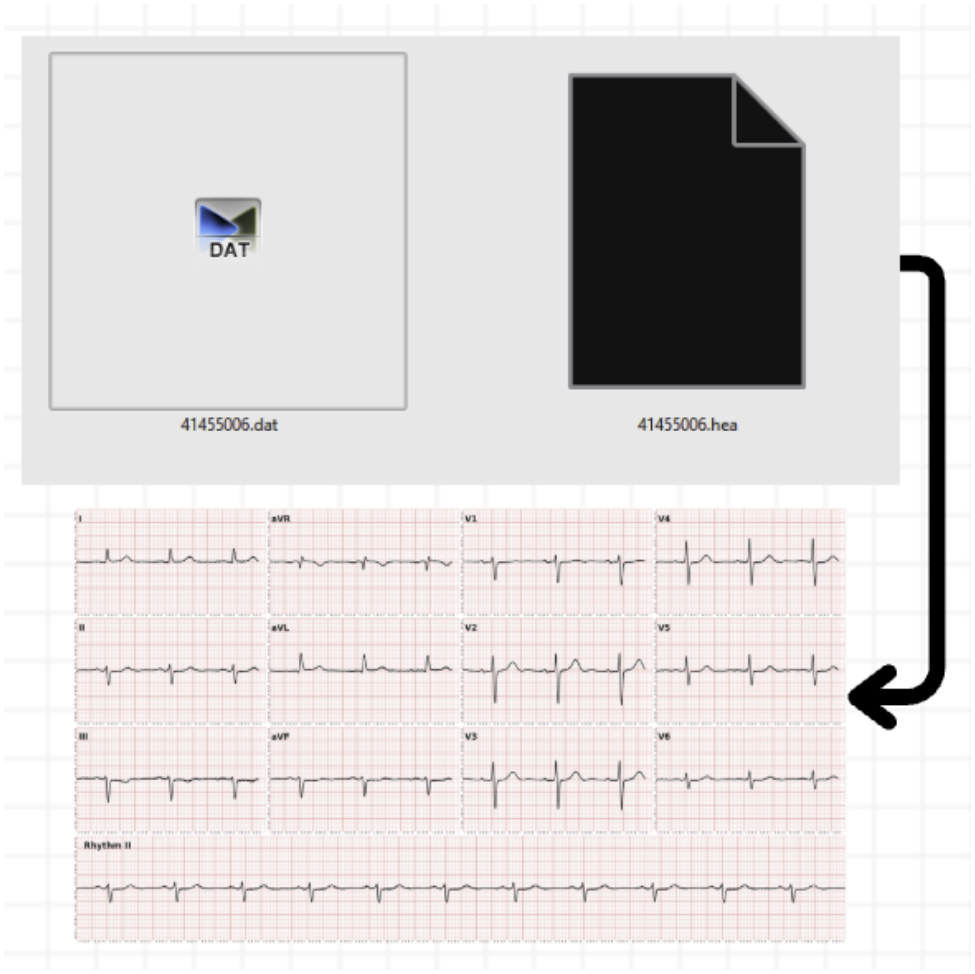
Conversion of raw binary waveform files into standardized 3×4 ECG images.

At inference time, user-uploaded photos or screenshots have alpha transparency flattened onto a solid white background, padded by an 80-pixel white border, and cropped via a YOLOv11s ECG detector (returning HTTP 400 if no valid ECG is detected to prevent hallucinations on non-ECG images). Scans pass through a mandatory 23-block RRDBNet Real-ESRGAN module (4x upscaling, 500-pixel tiling, *tile_pad* = 10) to sharpen fine waveform deflections.

### C. Structured JSON Clinical Target Construction

For open-vocabulary supervision, MIMIC-IV-ECG report fields (*report_0–report_17*) are cleaned and joined with “ | “ delimiters, while PTB-XL cardiologist reports and SCP summaries are paired with patient demographics (age, biological sex, and weight). Each sample is formatted into a deterministic instruction prompt requiring strict UTF-8 JSON output with two keys: “*report*” (concise clinical interpretation) and “*clinical_description*” (pathophysiological explanation) capped at 350 tokens. During causal language-model training, prompt, padding, and prepended visual token positions are masked with an ignore index of -100 so cross-entropy loss optimizes exclusively over the target JSON tokens:

## 4. SYSTEM ARCHITECTURE AND METHODOLOGY

### A. Multimodal Vision-Language Architecture

LifeLine.ai pairs a fine-tuned MedGemma-4B language backbone [4] with a visual encoder to translate 12-lead ECG plots into open-vocabulary clinical narratives (Fig. 4). We evaluated two vision-to-language fusion architectures under identical clinical supervision.

**Fig 3.**
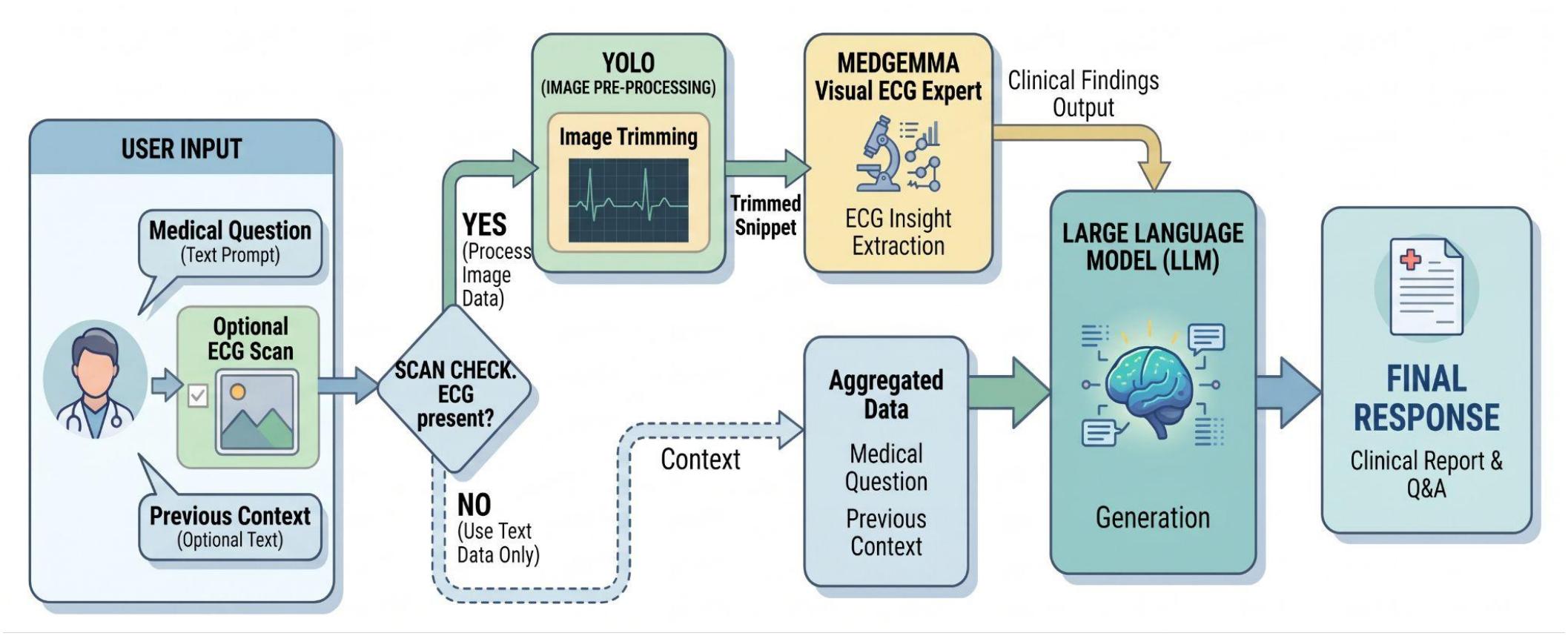
LifeLine.ai multimodal inference and dynamic Q&A pipeline.

**Fig 4.**
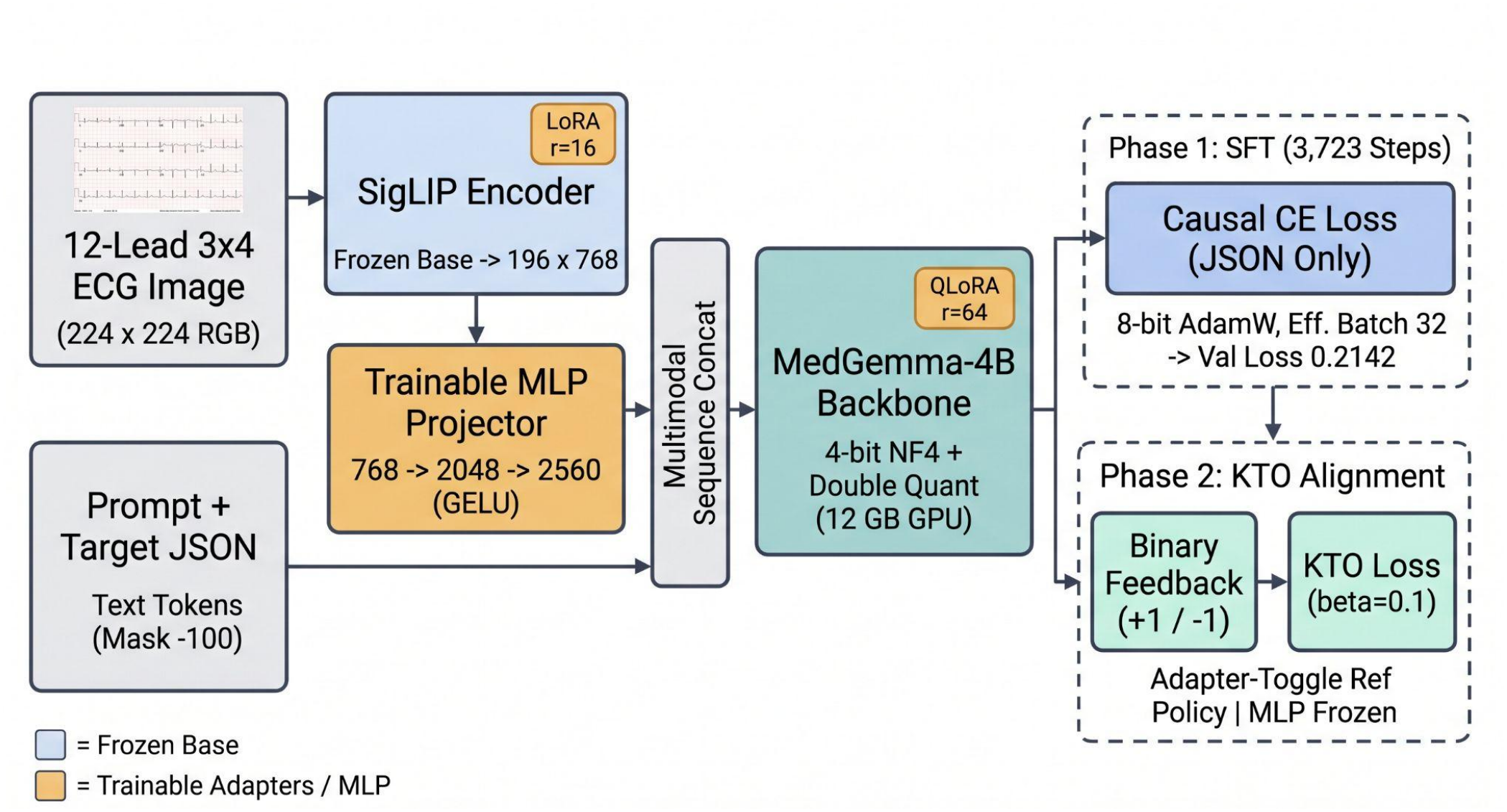
End-to-end SigLIP-MedGemma-4B QLoRA supervised fine-tuning (SFT) and post-SFT KTO preference-alignment architecture.

Our baseline paired a hierarchical Swin Transformer (swin-base-patch4-window7-224, 1024-dimensional output) [5] over 13 lead views (224 x 224 RGB) with an 8-head global-context attention module pre-trained across 71 SCP classes using Asymmetric Loss (*gamma_neg* = 4, *gamma_pos* = 1, *margin* = 0.05) [11]. Per-view features were compressed to 312 dimensions, flattened into static 4056-dimensional L2-normalized vectors, and projected into MedGemma-4B through a two-layer Multi-Layer Perceptron (MLP) with LayerNorm and GELU activations. However, shifted-window partitioning fragmented continuous horizontal waveforms and static pre-extracted features left a vision-language modality gap, plateauing validation loss near 0.25.

Our final architecture couples Google SigLIP (siglip-base-patch16-224) [6] end-to-end with MedGemma-4B (2560-dimensional hidden space). Base SigLIP weights remain frozen while Low-Rank Adaptation (LoRA) adapters (*r* = 16, *alpha* = 32, zero bias) [12] adapt q_proj and v_proj layers with gradient checkpointing. SigLIP produces 196 global visual tokens of 768 dimensions, mapped into MedGemma’s embedding space via a trainable float32 MLP projector: LayerNorm(768) → Linear(768, 2048) → GELU → Linear(2048, 2560) → LayerNorm(2560). Base MedGemma-4B weights are quantized to 4-bit NormalFloat (NF4) with double quantization and adapted via QLoRA (*r* = 64, *alpha* = 128) [7] on q_proj, k_proj, v_proj, and o_proj. The 196 projected visual tokens are prepended to embedded text tokens with visual attention masks set to 1 and visual labels masked to -100.

### B. Single-GPU QLoRA Supervised Fine-Tuning

Supervised fine-tuning (SFT) was designed for a single NVIDIA RTX A2000 GPU (12 GB VRAM). Four-bit NF4 quantization reduces base LLM memory consumption by approximately 75%, while non-reentrant gradient checkpointing prevents out-of-memory fragmentation. Given a micro-batch cap of 4 images, 8-step gradient accumulation simulates an effective *batch size* of 32. Optimization uses 8-bit AdamW (*weight decay* = 0.01, *max gradient norm* = 1.0) with a *cosine* learning rate schedule and 5% *linear warmup*, decaying from 9.95e-6 to 3.18e-8 over 3,723 global steps to preserve MedGemma’s medical pre-training. Masking visual, prompt, and padding tokens with -100 directs 100% of cross-entropy updates exclusively onto the target clinical JSON tokens.

### C. Post-SFT KTO Preference Alignment

To align generated reports with user expectations without costly paired preference datasets, we collected binary user feedback (+1.0 desirable, -1.0 undesirable) and built a separate Kahneman-Tversky Optimization (KTO) alignment pipeline [8] atop our SFT-trained SigLIP-MedGemma model. To avoid loading a duplicate 4B reference model into VRAM, reference log-probabilities (*log p_ref*) are computed in-place under *no_grad* by temporarily disabling vision and language LoRA adapters, while active policy log-probabilities (*log p_theta*) are computed with adapters enabled. Sequence log-probabilities are averaged over unmasked response tokens (*label* != -100), and the preference margin is optimized via a binary KTO objective:

L_KTO = -log sigma(y×beta×(log p_theta - log p_ref))

where y in {+1, -1} is the binary feedback label and beta = 0.1. During alignment, the MLP projector remains frozen for cross-modal stability while LoRA adapters update with 8-bit AdamW (learning rate 1e-6 for SigLIP, 5e-6 for MedGemma), 200 *warmup steps, cosine* decay, *bfloat16* precision, effective batch size 32 (8 × 4), and validation checkpointing every 300 steps.

### D. Inference Pipeline and Cloud Deployment Architecture

At inference time, uploaded ECG scans pass through two preprocessing stages before VLM reasoning (Fig. 3). First, low-resolution scans are enhanced with Real-ESRGAN (RealESRGAN_x4plus, 23-block RRDBNet, 64 feature channels, 32 growth channels) [13], applying 4x super-resolution with 500-pixel tiling (tile_pad = 10) in FP32 precision. Second, YOLOv11s [14] localizes the ECG grid after EXIF correction, RGBA-to-white flattening, and 80-pixel white padding (*confidence threshold* = 0.80, with unpadded fallback). Bounding boxes are de-padded to crop native-resolution waveforms, while non-ECG uploads trigger an HTTP 400 rejection.

Cropped ECGs are dispatched asynchronously (120-second timeout) to our fine-tuned MedGemma-4B VLM for structured clinical interpretation. Outputs are enriched by a reference summarizer (GPT-4.1-mini, *temperature* = 0.1, covering condition description, immediate response, age risk profile, and referral urgency) and synthesized by a consensus LLM on Groq (GPT-OSS-20B, temperature = 0.2). A dynamic Q&A route supports optional patient history, bypassing visual stages for text-only queries while invoking YOLO localization and MedGemma when an ECG image is attached.

LifeLine.ai is deployed across four decoupled cloud tiers (Fig. 5): (1) a Client Layer comprising a Vercel web application and native iOS/Android mobile apps with location-based cardiologist discovery; (2) a Middleware and Data Layer on Amazon EC2 running FastAPI, Firebase persistence, developer API key management, and the open-source lifelinecg-sdk Python library (v0.1.5); (3) an AI Application Layer hosted on Hugging Face exposing authenticated FastAPI endpoints; and (4) a Compute Layer executing MedGemma-4B inference on Modal serverless GPUs.

**Fig 5.**
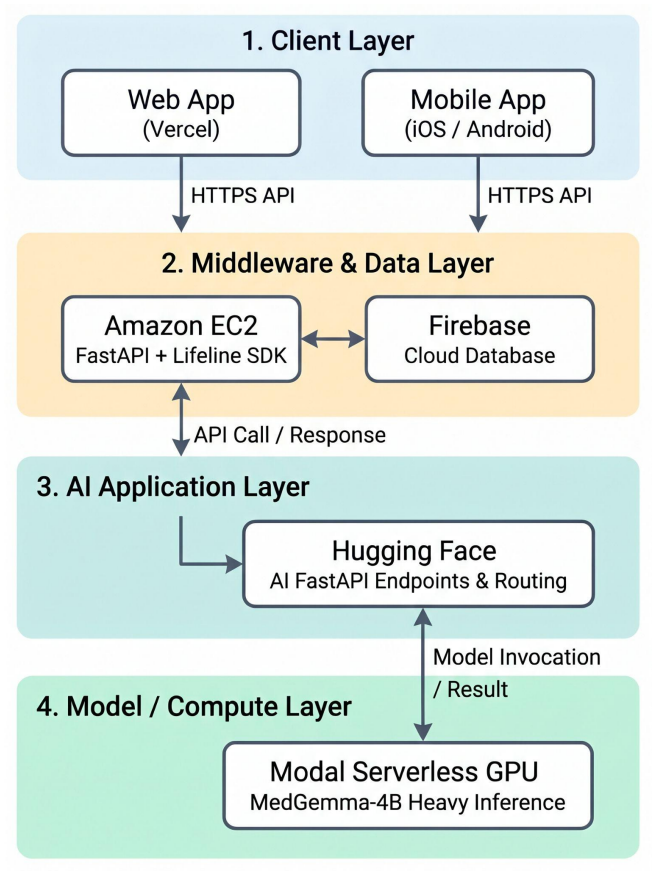
Four-tier LifeLine.ai cloud deployment architecture.

## 5. EXPERIMENTS AND RESULTS

### A. Evaluation Protocol and Metrics

Experiments were conducted on a single NVIDIA RTX A2000 GPU (12 GB VRAM) using 4-bit NormalFloat quantization, 8-bit AdamW, and an effective batch size of Validation performance was tracked on random hold-out splits: 97% - 3% (Stage 1) and 95% - 5% (Stage 2) for the Swin Transformer baseline, and 97.5% - 2.5% (seed 42, evaluated every 300 steps) for SigLIP-MedGemma. Prior to language alignment, Swin embeddings were audited via inter-patient cosine similarity, Euclidean distance, and Manhattan distance to verify class separation (cosine similarity below 0.50 across distinct pathologies, confirming absence of representation collapse).

Because LifeLine.ai performs open-ended clinical report generation rather than closed-set classification, diagnostic performance was evaluated using Open-Vocabulary Precision and Contextual F1 Score across generated clinical narratives and interactive question-answering (Q&A) outputs.

### B. Vision Encoder Ablation: SigLIP vs. Swin Transformer

We compared the Swin Transformer baseline against our end-to-end LoRA-adapted Google SigLIP encoder paired with MedGemma-4B (Fig. 4). Over approximately 9,800 global steps, Swin-MedGemma reduced validation loss from 0.51 to ∼0.32 (step 2,000) and ∼0.28 (step 4,000), plateauing near 0.25 from step 6,000 onward while smoothed training loss drifted toward 0.20–0.21. This generalization floor reflects horizontal waveform fragmentation caused by shifted-window partitioning and static 4056-dimensional pre-extracted features.

In contrast, coupling SigLIP with MedGemma-4B through our trainable 768-to-2560 MLP projector produced faster, lower-variance convergence under cosine decay from 9.95e-6 to 3.18e-8 (Fig. 6). Across 3,723 global steps, training loss decreased from 1.0882 (step 300) to 0.5142 (step 1,241), 0.2685 (step 1,500), and 0.2033 (step 3,723).

**Fig 6.**
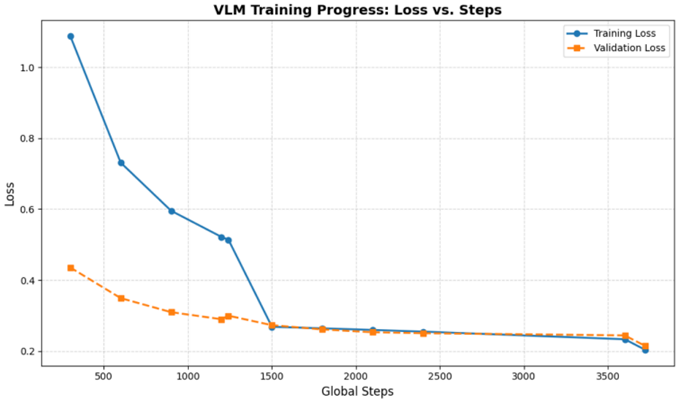
Training and validation loss curves for SigLIP-MedGemma-4B across 3,723 global steps.

**Fig 7.**
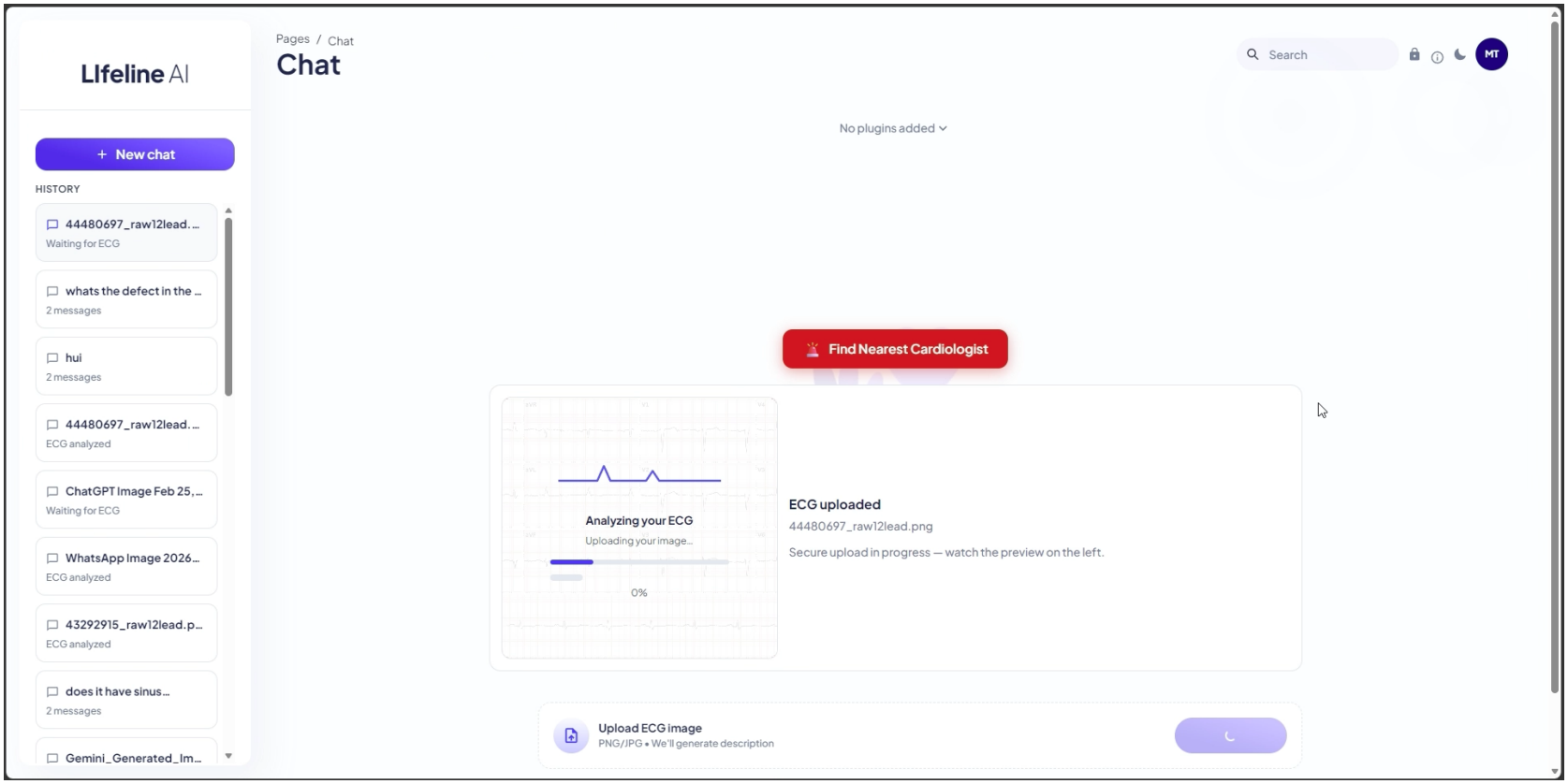
LifeLine.ai web chat interface initiating ECG image upload and preprocessing, displaying the upload preview panel, session history sidebar, PNG/JPG submission bar, and the emergency nearest-cardiologist action button.

Validation loss declined steadily from 0.4357 (step 300) to 0.2898 (step 1,200), 0.2730 (step 1,500), 0.2504 (step 2,400), 0.2442 (step 3,600), and 0.2142 (∼0.21) at step 3,723, outperforming Swin by ∼0.036 validation loss in fewer than 40% of the training steps with virtually no train-validation gap.

**Fig 8.**
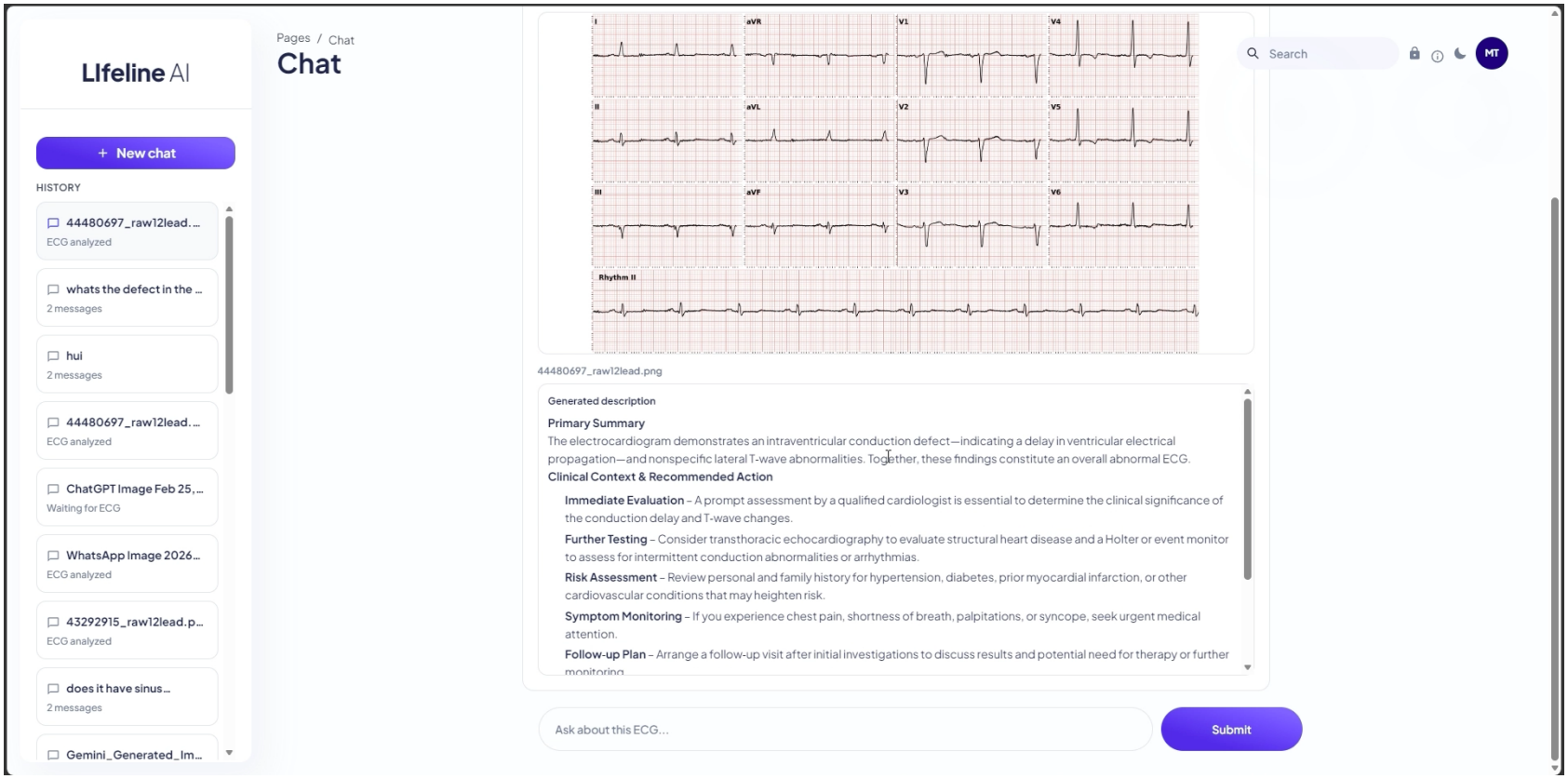
Automated ECG diagnostic report view displaying an uploaded 12-lead ECG chart with a continuous Lead II rhythm strip above the structured Primary Summary and Clinical Context & Recommended Action sections (Immediate Evaluation, Further Testing, Risk Assessment, Symptom Monitoring, and Follow-up Plan), paired with the follow-up query bar.

**Fig 9.**
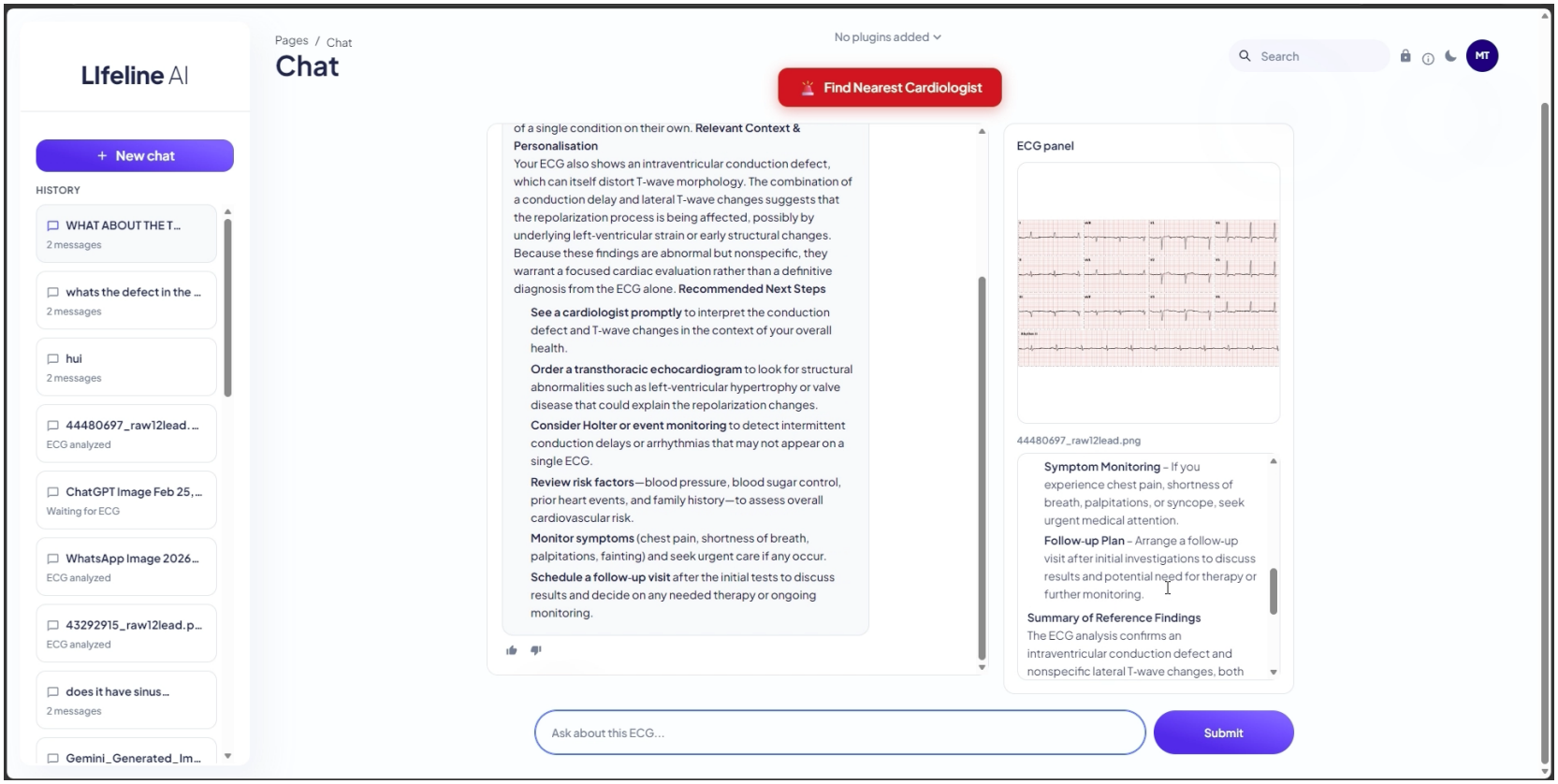
Interactive clinical Q&A split-panel interface presenting contextualized follow-up guidance (Relevant Context & Personalisation and Recommended Next Steps) with binary thumbs-up/thumbs-down user feedback controls supporting KTO alignment on the left, alongside an anchored ECG reference panel on the right.

**Fig 10.**
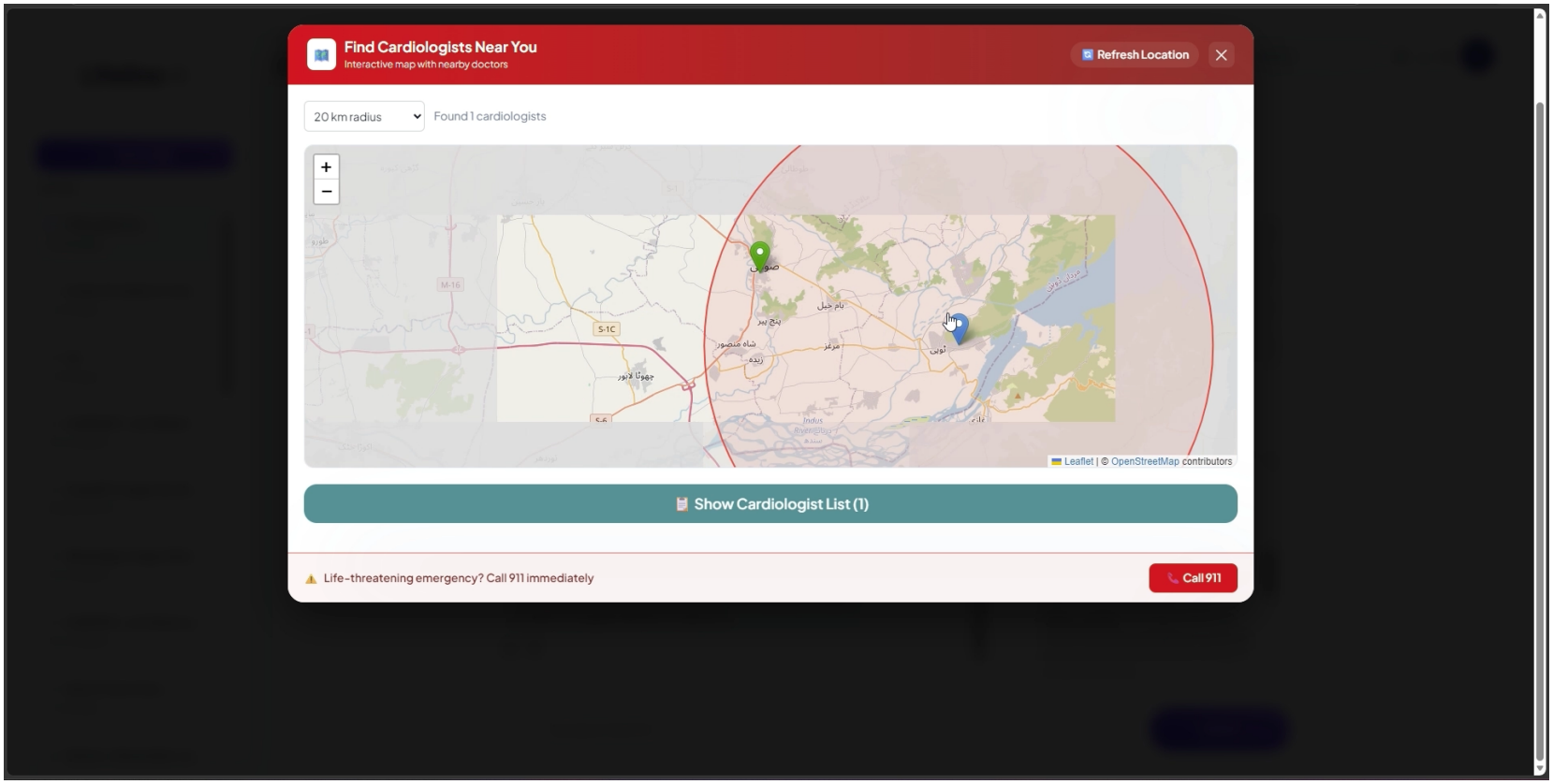
Emergency cardiologist discovery modal powered by Leaflet and OpenStreetMap, showing a configurable 20 km search radius, geographic markers for user and specialist locations, a cardiologist list drawer, and an emergency call shortcut.

**Fig 11.**
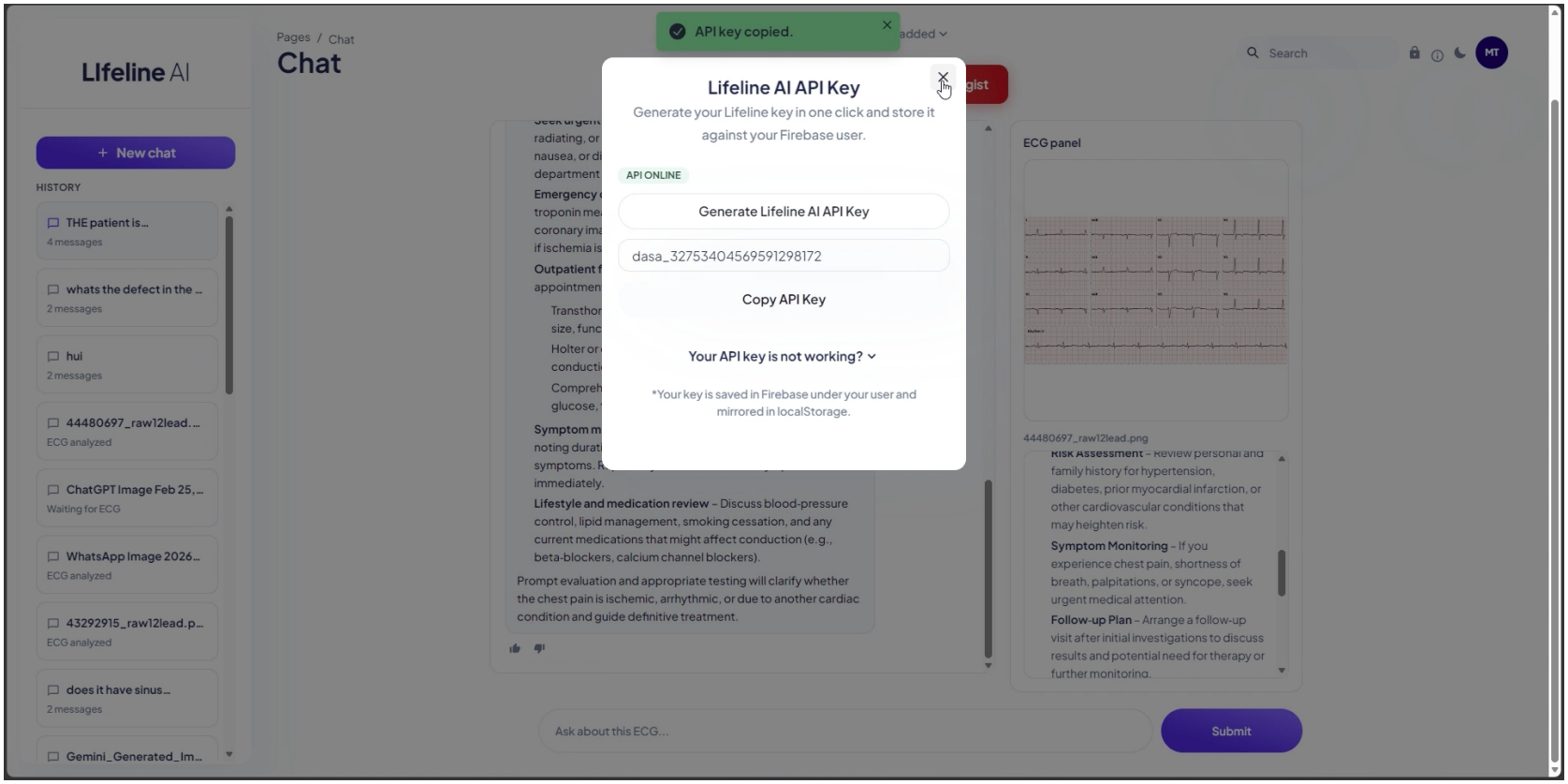
Developer API key provisioning modal displaying live API availability, one-click generation and clipboard copying of a dasa_-prefixed credential, and Firebase account synchronization status.

### C. Comparison with Prior ECG Analysis Techniques

Tables 2 and 3 compare LifeLine.ai against ECG-Image [1], Digitized-ECG [2], and xECGArch [3].

**Table 1.** REPRESENTATIVE NORMAL AND ABNORMAL STRUCTURED JSON CLINICAL TARGETS.

| Case Type | Report | Clinical Description |
|---|---|---|
| Normal / Benign | Sinus bradycardia otherwise normal ECG | Heart rate is slower than normal, with electrical signals originating regularly from the sinus node. |
| Pathological / Abnormal | Fast atrial fibrillation, left axis | Provided image depicts atrial fibrillation. This means the heart's upper chambers are beating irregularly and quickly, without a clear pattern. The heart's lower chambers are also beating quickly, which can make you feel tired and weak. Atrial fibrillation can be dangerous because it can lead to blood clots that may cause a stroke. |

**Table 2.**
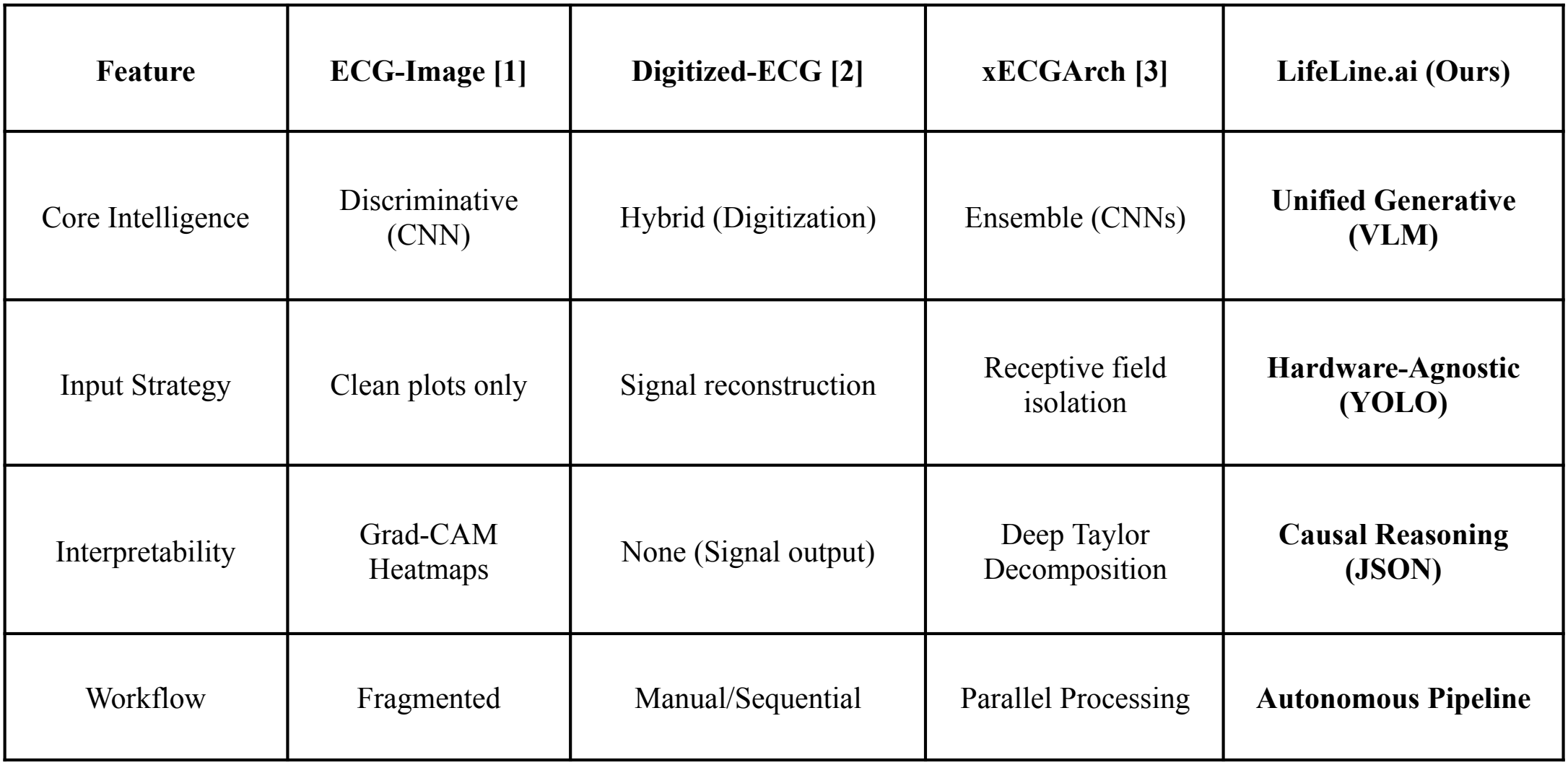
ARCHITECTURAL AND WORKFLOW COMPARISON WITH PRIOR ECG TECHNIQUES.

**Table 3.**
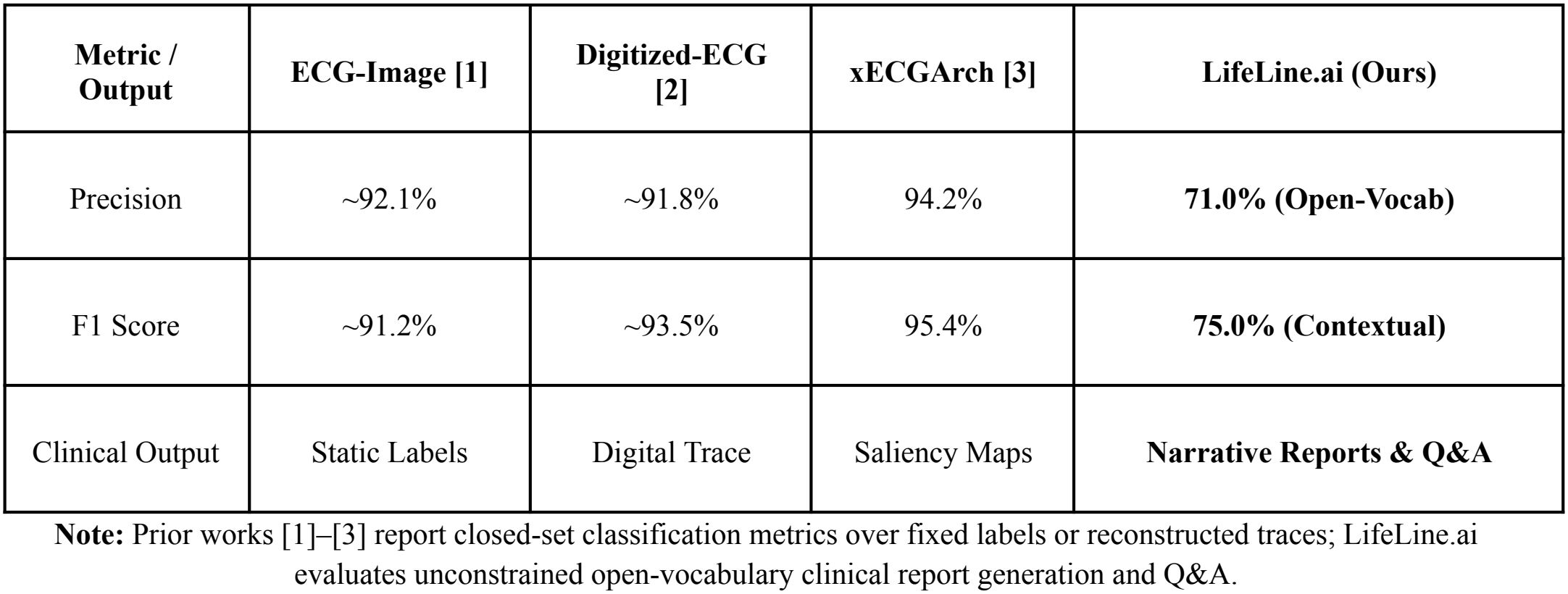
QUANTITATIVE BENCHMARK AND CLINICAL OUTPUT COMPARISON.

Direct numerical comparison requires distinguishing task formulation. Prior baselines achieve 91.2%–95.4% precision and F1 on closed-set classification over predefined taxonomies. LifeLine.ai instead addresses unconstrained open-vocabulary report generation and interactive Q&A from raw images using a compact 50,000-sample subset; its 71.0% open-vocabulary precision and 75.0% contextual F1 therefore reflect free-form clinical reasoning rather than closed-library discrimination.

### D. Qualitative Output and Inference Analysis

End-to-end inference evaluation confirms that upstream visual guards enforce safety prior to clinical generation. Non-ECG uploads trigger an immediate HTTP 400 rejection from YOLOv11s, preventing hallucinated reports on out-of-domain images. Valid ECG crops are processed asynchronously by MedGemma-4B (120-second timeout) to produce structured JSON findings, which are refined by clinical summarization (GPT-4.1-mini, *temperature* = 0.1) and consensus synthesis (GPT-OSS-20B, *temperature* = 0.2) into four clinical sections: condition description, immediate response, age risk profile, and referral urgency. Text-only follow-up queries with patient history bypass visual segmentation and VLM inference directly. Automated integration tests across the backend, SDK, and client layers verify request authentication, YOLO non-ECG rejection, malformed-JSON fallbacks, and modality routing. Two hardware-driven constraints remain: resizing 12-lead grids to 224 x 224 pixels can attenuate low-amplitude P-waves, and serverless GPU cold starts bound worst-case latency.

## 6. LIMITATIONS, ERROR ANALYSIS, AND ETHICAL CONSIDERATIONS

### A. Compute, Data Scale, and Resolution Constraints

All supervised fine-tuning and alignment experiments were conducted under strict single-GPU academic hardware constraints on an NVIDIA RTX A2000 GPU (12 GB VRAM). Because of memory, storage, and training-time limits, the final multimodal VLM was trained on a compact, class-balanced subset of approximately 50,000 ECG records, representing roughly 6.25% of the >800,000 raw recordings available across PTB-XL [9] and MIMIC-IV-ECG [10], using 4-bit NF4 quantization, a micro-batch size of 4, 224 x 224 RGB image resizing, and a capped training budget of 3,723 global steps. Because validation loss was still declining at the end of training (reaching 0.2142) alongside 75.0% contextual F1 on this limited ∼50,000-sample regime, training for longer schedules (additional epochs and global steps) while scaling to the full multi-hospital corpus with larger batch sizes and higher input resolution is expected to yield substantial gains in diagnostic precision and rare-arrhythmia recall.

### B. Error Analysis

Three primary failure modes emerge under these resource constraints. First, limiting training to ∼50,000 samples and 3,723 steps restricts exposure to rare, long-tail cardiac pathologies and complex multi-condition co-occurrences. Second, compressing twelve standard leads and a continuous rhythm strip into 224 × 224 pixels can attenuate low-amplitude morphological cues such as subtle P-waves or borderline ST-segment deviations. Third, open-vocabulary evaluation relies on automated factual and contextual matching rather than blinded cardiologist grading, where valid medical synonyms can depress strict precision scores while subtle clinical omissions require expert review; in cloud deployment, serverless GPU cold starts also bound worst-case inference latency.

### C. Ethical and Clinical Safety Considerations

LifeLine.ai is strictly a research prototype and clinical decision-support aid, it is not a regulatory-cleared medical device and must not be used for standalone clinical diagnosis or emergency triage without supervision by a qualified physician. All training data were derived from de-identified public PhysioNet benchmarks (PTB-XL [9] and MIMIC-IV-ECG [10]). Any real-world clinical deployment requires prospective multi-center validation, institutional review, and continuous clinician-in-the-loop oversight to mitigate generative hallucination risks.

## 7. CONCLUSION AND FUTURE WORK

We presented LifeLine.ai, an end-to-end multimodal Vision-ECG-Language system for open-vocabulary 12-lead ECG interpretation, structured JSON clinical reporting, and interactive medical Q&A trained on a single 12 GB GPU. By synthesizing standardized 3×4 ECG grids from PhysioNet waveforms and coupling a LoRA-adapted Google SigLIP encoder with a 4-bit quantized MedGemma-4B backbone via a trainable MLP projector, our model reduced validation loss to 0.2142 in 3,723 steps, outperforming a windowed Swin Transformer baseline (∼0.25 at 9,800 steps) while achieving 71.0% open-vocabulary precision and 75.0% contextual F1 using only ∼50,000 training records. Combined with a standalone KTO user-feedback alignment stage, Real-ESRGAN/YOLOv11s preprocessing, and a four-tier cloud deployment with the lifelinecg-sdk package, LifeLine.ai demonstrates a practical pathway beyond static closed-set ECG classifiers. Future work will focus on scaling training from ∼50,000 records to the full >800,000-record PhysioNet archive on multi-GPU clusters, adopting higher-resolution vision encoders to preserve fine P-wave and ST morphology, and conducting blinded evaluations with board-certified cardiologists.

## Data Availability

All ECG datasets analyzed in this study are publicly available de-identified repositories hosted on PhysioNet: PTB-XL (https://physionet.org/content/ptb-xl/) and MIMIC-IV-ECG (https://physionet.org/content/mimic-iv-ecg/1.0/). The source code and preprocessing scripts for LifeLine.ai are publicly available at https://github.com/Asad939asad/LifeLine.ai-.

https://physionet.org/content/ptb-xl/

https://physionet.org/content/mimic-iv-ecg/1.0/

## APPENDIX

### SOFTWARE IMPLEMENTATION AND USER INTERFACE

#### A. System Stack and API Endpoints

LifeLine.ai is implemented as a four-tier cloud architecture separating user interaction, middleware persistence, AI orchestration, and serverless GPU execution. The Client Layer comprises a Vercel-hosted web application and native iOS and Android mobile clients communicating over HTTPS with an Amazon EC2 FastAPI middleware server. User authentication, chat logs, and developer credentials are persisted in Firebase, while API key validation is backed by a lightweight SQLite registry (dasa_ keys capped at 2 active keys per email). The middleware invokes the Hugging Face-hosted AI Application Layer (Lifeline ECG Vision API), which orchestrates YOLOv11s segmentation, asynchronous MedGemma-4B inference on Modal serverless GPUs, and downstream consensus summarization.

Five RESTful FastAPI endpoints expose the core service:

- GET /: Returns service readiness status.
- POST /v1/keys/generate: Issues a 20-digit dasa_ API key authorized via the admin-secret header.
- DELETE /v1/keys/delete-oldest: Revokes the oldest active key registered to an email address.
- POST /v1/analyze: Authenticated via x-api-key; accepts a multipart ECG upload, runs YOLOv11s cropping, MedGemma-4B interpretation, and consensus synthesis, returning status, pipeline_metrics (yolo_crop_size), and final_report.
- POST /v1/analyze-dynamic: Authenticated via x-api-key; accepts a required prompt, optional patient context, and optional ECG upload, returning modality_used (Text + Image or Text Only) and final_report.

#### B. Python SDK Usage (lifelinecg-sdk)

To simplify clinical integration, the client interface is distributed as the lifelinecg-sdk Python package (v0.1.5). Developers initialize LifelineClient with an active API key to invoke static diagnosis (analyze) or context-aware multimodal Q&A (analyze_dynamic), alongside administrative key methods (*generate_api_key, delete_oldest_api_key*):

from lifeline import LifelineClient

client = LifelineClient(api_key=“dasa_your_active_key”)

static_res = client.analyze(image_path=“sample_12lead_ecg.png”)

qa_res = client.analyze_dynamic(prompt=“Evaluate the lateral T-wave changes.”, image_path=“sample_12lead_ecg.png”, context=“65-year-old male presenting with chest discomfort.”)

print(static_res[“final_report”], qa_res[“modality_used”])

#### C. Application Interface Previews

## REFERENCES

[1] R. Ao and G. He, “Image based deep learning in 12-lead ECG diagnosis,” Frontiers in Artificial Intelligence, vol. 5, art. 1087370, Jan. 2023, doi: 10.3389/frai.2022.1087370.

[2] S. Sattar et al., “Cardiac arrhythmia classification using advanced deep learning techniques on digitized ECG datasets,” Sensors, vol. 24, no. 8, art. 2484, Apr. 2024, doi: 10.3390/s24082484.

[3] M. Goettling, A. Hammer, H. Malberg, and M. Schmidt, “xECGArch: A trustworthy deep learning architecture for interpretable ECG analysis considering short-term and long-term features,” Scientific Reports, vol. 14, no. 1, art. 13122, Jun. 2024, doi: 10.1038/s41598-024-63656-x.

[4] A. Sellergren et al., “MedGemma Technical Report,” arXiv preprint arXiv:2507.05201, Jul. 2025.

[5] Z. Liu, Y. Lin, Y. Cao, H. Hu, Y. Wei, Z. Zhang, S. Lin, and B. Guo, “Swin Transformer: Hierarchical vision transformer using shifted windows,” in Proc. IEEE/CVF Int. Conf. Comput. Vis. (ICCV), Oct. 2021, pp. 10012–10022.

[6] X. Zhai, B. Mustafa, A. Kolesnikov, and L. Beyer, “Sigmoid loss for language image pre-training,” in Proc. IEEE/CVF Int. Conf. Comput. Vis. (ICCV), Oct. 2023, pp. 11975–11986.

[7] T. Dettmers, A. Pagnoni, A. Holtzman, and L. Zettlemoyer, “QLoRA: Efficient finetuning of quantized LLMs,” in Adv. Neural Inf. Process. Syst. (NeurIPS), vol. 36, Dec. 2023, pp. 10088–10115.

[8] K. Ethayarajh, W. Xu, N. Muennighoff, D. Jurafsky, and D. Kiela, “KTO: Model alignment as prospect theoretic optimization,” in Proc. Int. Conf. Mach. Learn. (ICML), Jul. 2024, pp. 12650–12668.

[9] P. Wagner, N. Strodthoff, R.-D. Bousseljot, D. Kreiseler, F. I. Lunze, W. Samek, and T. Schaeffter, “PTB-XL, a large publicly available electrocardiography dataset,” Scientific Data, vol. 7, no. 1, art. 154, May 2020, doi: 10.1038/s41597-020-0495-6.

[10] B. Gow et al., “MIMIC-IV-ECG: Diagnostic Electrocardiogram Matched Subset,” version 1.0, PhysioNet, 2023, doi: 10.13026/4nqg-sb35.

[11] T. Ridnik, E. Ben-Baruch, N. Zamir, A. Noy, I. Friedman, M. Protter, and L. Zelnik-Manor, “Asymmetric loss for multi-label classification,” in Proc. IEEE/CVF Int. Conf. Comput. Vis. (ICCV), Oct. 2021, pp. 82–91.

[12] E. J. Hu et al., “LoRA: Low-rank adaptation of large language models,” in Proc. Int. Conf. Learn. Represent. (ICLR), Apr. 2022.

[13] X. Wang, L. Xie, C. Dong, and Y. Shan, “Real-ESRGAN: Training real-world blind super-resolution with pure synthetic data,” in Proc. IEEE/CVF Int. Conf. Comput. Vis. Workshops (ICCVW), Oct. 2021, pp. 1905–1914.

[14] G. Jocher and J. Qiu, “Ultralytics YOLO11,” version 11.0.0, Ultralytics, 2024. [Online]. Available: https://github.com/ultralytics/ultralytics

[15] A. H. Ribeiro, M. H. Ribeiro, G. M. M. Paixão, D. M. Oliveira, P. R. Gomes, J. A. Canazart, M. P. S. Ferreira, C. R. Andersson, P. W. Macfarlane, W. Meira Jr., T. B. Schön, and A. L. P. Ribeiro, “Automatic diagnosis of the 12-lead ECG using a deep neural network,” Nature Communications, vol. 11, no. 1, art. 1760, Apr. 2020, doi: 10.1038/s41467-020-15432-4.

[16] V. Sangha et al., “Automated multilabel diagnosis on electrocardiographic images and signals,” Nature Communications, vol. 13, no. 1, art. 1583, Mar. 2022, doi: 10.1038/s41467-022-29153-3.

[17] K. K. Shivashankara, Deepanshi, A. M. Shervedani, M. A. Reyna, G. D. Clifford, and R. Sameni, “ECG-Image-Kit: A synthetic image generation toolbox to facilitate deep learning-based electrocardiogram digitization,” Physiological Measurement, vol. 45, art. 055019, May 2024, doi: 10.1088/1361-6579/ad4954.

[18] T. Tu et al., “Towards generalist biomedical AI,” NEJM AI, vol. 1, no. 3, Feb. 2024, doi: 10.1056/AIoa2300138.

[19] S. Zhang et al., “BiomedCLIP: A multimodal biomedical foundation model pretrained from fifteen million scientific image-text pairs,” arXiv preprint arXiv:2303.00915, Mar. 2023.

[20] R. Liu, Y. Bai, X. Yue, and P. Zhang, “Teach multimodal LLMs to comprehend electrocardiographic images,” arXiv preprint arXiv:2410.19008, Oct. 2024.

[21] L. Ouyang et al., “Training language models to follow instructions with human feedback,” in Adv. Neural Inf. Process. Syst. (NeurIPS), vol. 35, Dec. 2022, pp. 27730–27744.

[22] R. Rafailov, A. Sharma, E. Mitchell, S. Ermon, C. D. Manning, and C. Finn, “Direct preference optimization: Your language model is secretly a reward model,” in Adv. Neural Inf. Process. Syst. (NeurIPS), vol. 36, Dec. 2023, pp. 53728–53741.

